# Plasma ceramides as a sexually dimorphic biomarker of pancreatic cancer-induced cachexia

**DOI:** 10.1101/2020.06.01.20111492

**Authors:** Jeffery M. Chakedis, Mary E. Dillhoff, Carl R. Schmidt, Priyani V. Rajasekera, David C. Evans, Terence M. Williams, Denis C. Guttridge, Erin E. Talbert

**Author notes:** Present Address: Department of General Surgery, The Permanente Medical Group, Kaiser Permanente Walnut Creek Medical Center, CA 94596. Present Address: Department of Surgery, West Virginia University, Morgantown, WV, 26506. Present Address: OhioHealth Trauma Services, Columbus, OH, 43215. Present Address: Department of Health and Human Physiology and Holden Comprehensive Cancer Center, University of Iowa, Iowa City, IA, 52242. Correspondence to Denis C. Guttridge and Erin E. Talbert.

## Abstract

**Background:** Cancer patients who lose weight have low treatment tolerance and poor outcomes compared to cancer patients without weight loss, termed cachexia. Despite the clear increased risk for patients, diagnosing cachexia still primarily relies on self-reported weight loss. A reliable biomarker to identify patients with cancer cachexia would be a valuable tool to improve clinical decision making and identification of patients at risk of adverse outcomes.

**Methods:** Targeted metabolomics, including panels of amino acids, tricarboxylic acids, fatty acids, acylcarnitines, and sphingolipids, were conducted on plasma samples from patients with confirmed pancreatic ductal adenocarcinoma (PDAC) with and without cachexia and control patients without cancer. Receiver Operating Characteristic (ROC) analysis was undertaken to establish if any metabolite could effectively serve as a biomarker of cachexia.

**Results:** Targeted profiling revealed that cachectic patients had decreased circulating levels of three sphingolipids compared to either non-cachectic PDAC patients or patients without cancer. The ratio of C18-ceramide to C24-ceramide (C18:C24) outperformed a number of other previously proposed biomarkers of cachexia (area under ROC = 0.810). It was notable that some biomarkers, including C18:C24, were only elevated in cachectic males.

**Conclusion:** Our findings identify C18:C24 as a potentially new biomarker of PDAC-induced cachexia that also highlight a previously unappreciated sexual dimorphism in cancer cachexia.

**Trial registration:** None.

**Funding:** Support was provided through a pilot grant from U24DK100469 from the National Institute of Diabetes and Digestive and Kidney Diseases (The Mayo Clinic), National Cancer Institute P30CA016058 (The Ohio State University), National Cancer Institute R01CA180057 (DCG), American Cancer Society PF-15-156-01-CSM (EET), and a Weiss Postdoctoral Fellowship (EET).

## Introduction

Cachexia is a common clinical syndrome of cancer patients and is characterized by involuntary weight loss due to depletion of skeletal muscle and adipose tissue (1). Although anorexia is a component of the cachexia syndrome, supplemental nutrition is generally ineffective at preventing or reversing weight loss (2, 3). Patients with cachexia have higher morbidity and mortality, due in part to decreased tolerance of chemo- and radiotherapy and worse surgical outcomes (4-8). To date, there remains no effective treatment for cachexia in cancer patients.

Pancreatic ductal adenocarcinoma (PDAC) patients have amongst the highest incidence of cachexia, with estimates as high as 70% of patients affected (8-10). Cachexia also tends to be severe in PDAC patients, with body weight losses averaging ~14% of pre-illness weight (10). Furthermore, while an increased incidence of cachexia is associated with advanced disease, a significant proportion of PDAC patients already meet cachexia criteria at the time of cancer diagnosis (8, 11).

Although significant advances have been made in identifying some potential underlying mechanisms leading to muscle wasting and weight loss, cachexia remains surprising challenging to diagnose. Providers rely on often unreliable self-reported weight loss, likely leading to an underappreciation of the incidence of cachexia (12-14). Moreover, although an international consensus defines cachexia as a 5% loss of pre-illness body weight, recent studies in PDAC patients indicate that only greater losses may be associated with poor outcomes (8, 15). Therefore, there is significant need for a biomarker of cancer cachexia to more accurately identify this syndrome and identify patients at greater risk of morbidity and mortality.

In a recent study, we performed a multiplex analysis on a targeted panel of circulating cytokines, chemokines, and growth factors in early-stage PDAC patients in search of a potential biomarker for cachexia. We were surprised to find that a number of classical inflammatory cytokines that have been considered as drivers of cancer cachexia, including tumor necrosis factor (TNF), interleukin 1β (IL-1β), interleukin-6 (IL-6), and interferon-γ (IFN-γ) were not associated with weight loss (16). Given that cachexia is ultimately a metabolic syndrome, we took a different approach in this study using metabolomic profiling, which has been similarly examined by other investigators (17-22). Unlike these previous findings, our results revealed a general regulation of circulating sphingolipids in cachectic PDAC patients. Specifically, we identified a ceramide ratio that was associated with cachexia. In head-to-head comparisons, C18-ceramide to C24-ceramide (C18:C24) outperformed a number of circulating factors previously proposed as biomarkers of cancer cachexia. Furthermore, our results identified an unexpected sexual dimorphism in C18:C24.

## Results

### Experimental design

Patients undergoing an abdominal operation for suspected or confirmed PDAC or other benign conditions were eligible for enrollment into our Cancer Cachexia Tissue Registry (15, 16, 23, 24). Patients electing to participate were asked about their history of weight loss at time of their pre-operative clinic visit. When possible, weight loss data were confirmed by existing medical records.

Plasma samples from three groups of patients (n=10/group; 5 males and 5 females) were chosen for analysis from our biobank. These groups included: 1) Control patients without active cancer undergoing abdominal operations for a variety of diagnoses with no recent history of weight loss; 2) Weight-stable (non-cachectic) patients with histologically confirmed PDAC; and 3) PDAC patients with cachexia, defined as more than 5% weight loss over the previous 6-month period (1). Notably, at time of sample collection, PDAC cohorts were treatment-naïve. Clinical data from each group appear in Supplemental Table 1. To the best of our ability, patients were matched based on age and body mass index. PDAC patients with cachexia exhibited a mean weight loss of 10.3%. Plasma from these cohorts were screened by targeted metabolomic profiling for pre-established panels of amino compounds, tricarboxylic acids, fatty acids, acylcarnitines, and sphingolipids.

### Cachectic PDAC patients have altered plasma sphingolipid content

Following data normalization and correction for multiple hypothesis testing (false discovery rate <0.05), no significant differences were identified in the panels of amino compounds, fatty acids, or acylcarnitines between PDAC cachectic, non-cachectic, or control patients. In targeted profiling of tricarboxylic acid cycle metabolites, only fumarate was significantly decreased in non-cachectic PDAC patients compared to cachectic PDAC and control patients (data not shown).

With regards to targeted profiling of plasma sphingolipids, Sphingosine (SPH), Sphinganine (SPA),Sphingosine-1-phosphate (S1P), C14-Ceramide (C14), C16-Ceramide (C16), C18:1-Ceramide (C18-1), C18-Ceramide (C18, C20-Ceramide (C20), C22-Ceramide (C22), C24:1-Ceramide (C24-1), and C24-Ceramide (C24) were all detected in our plasma samples.C8-Ceramide was not detected in any samples and thus excluded from further analysis. Plasma from cachectic patients had a distinct sphingolipid signature compared to plasma from control and non-cachectic PDAC patients (Figure 1A). In particular, the concentrations of C22, C24, and S1P were decreased in cachectic patients compared to both other groups (Figure 1B, Supplemental Figure 1). Given these trends, we sought to confirm our findings by repeating our analysis in additional samples (clinical data for selected patients appears in Supplemental Table 2). Importantly, a similar pattern of plasma ceramides was observed in cachectic patients (Figure 2 and Supplemental Figure 2A).

**Figure 1.**
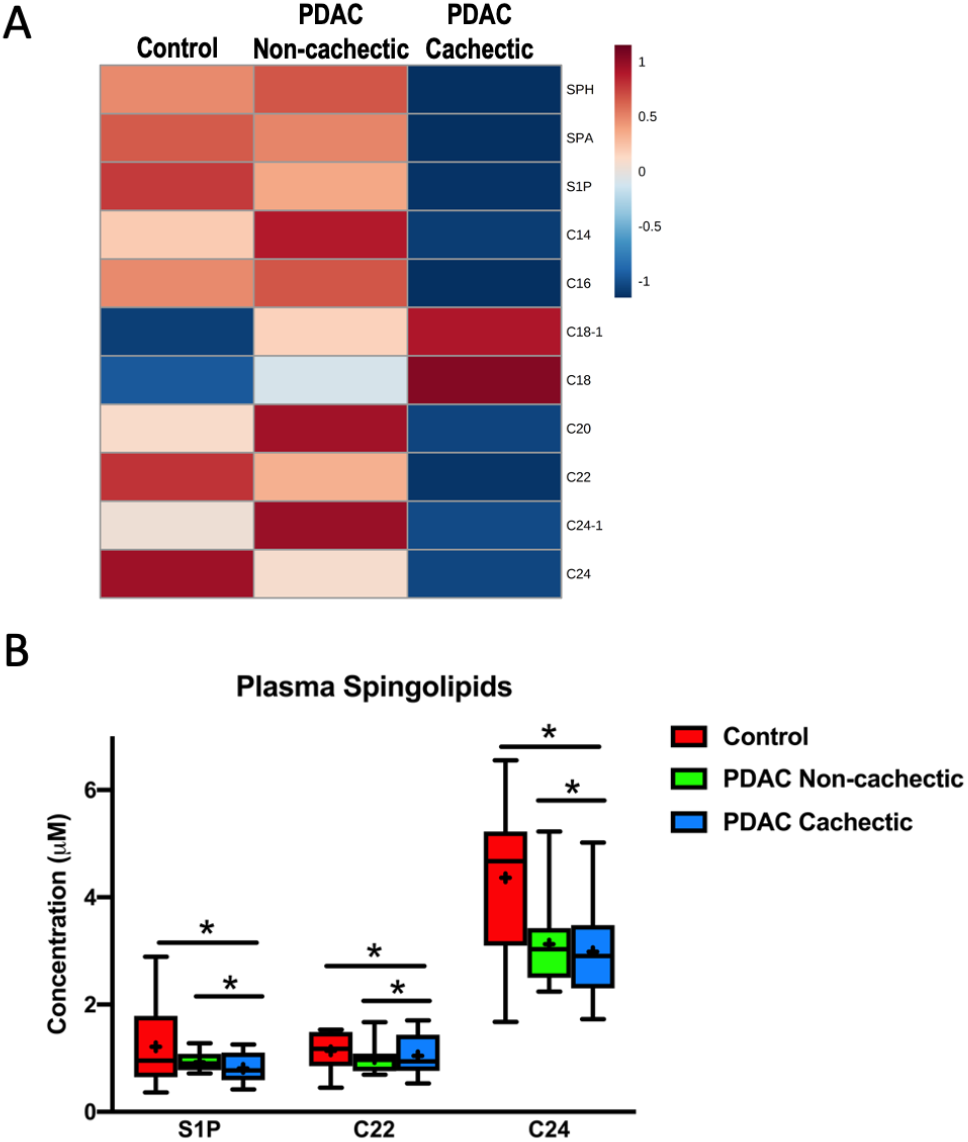
Plasma sphingolipid content is altered in cachectic PDAC patients. (A) Heatmap of plasma sphingolipids in control patients without cancer (n=10), noncachectic PDAC patients (n=10), and cachectic PDAC patients (n=10). The heatmap was generated in MetaboAnalyst using Euclidean distant measurements, auto-scaled by samples, and colored according to the normalized values. (B) Plasma levels of S1P, C22, and C24 are significantly decreased in cachectic PDAC patients compared to both control patients and non-cachectic PDAC patients. In the box-and-whiskers plot, the box is the 25^th^/75^th^ percentile with minimum-to-maximum whiskers. The line represents the median with the + representing the mean. * represents p < 0.05 by one-way ANOVA with Fisher’s LSD post-hoc using a false discovery rate of 0.05.

**Figure 2.**
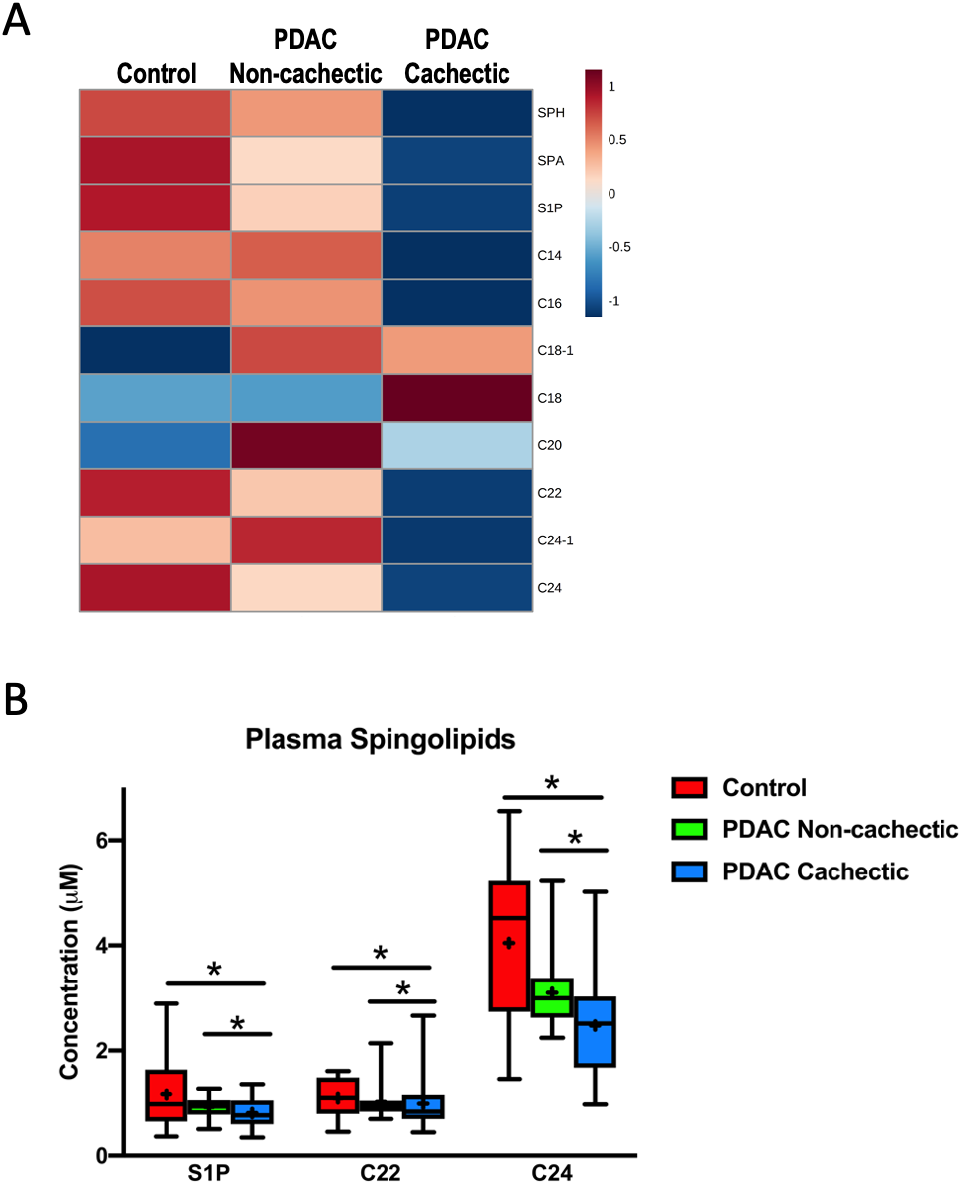
Plasma sphingolipid content is altered in an expanded cohort of cachectic PDAC patients. (A) Heatmap of plasma sphingolipids in control patients without cancer (n=14), non-cachectic PDAC patients (n=18), and cachectic PDAC patients (n=35). The heatmap was generated in MetaboAnalyst using Euclidean distant measurements, autoscaled by samples, and colored according to the normalized values. (B) Plasma levels of S1P, C22, and C24 are significantly decreased in cachectic PDAC patients compared to both control patients and non-cachectic PDAC patients. In the box-and-whiskers plot, the box is the 25^th^/75^th^ percentile with minimum-to-maximum whiskers. The line represents the median with the + representing the mean. * represents p < 0.05 by one-way ANOVA with Fisher’s LSD post-hoc using a false discovery rate of 0.05.

Because ceramide values are measured in comparison to known references and thus are quantitative, data from the first and second cohorts were collapsed into a single analysis (clinical data for the combined cohort appears in Table 1). Similar to our initial analysis, S1P, C22, and C24 remained significantly decreased in cachectic PDAC patients compared to non-cachectic PDAC patients and non-cancer controls (Figure 2 and Supplemental Figure 2B). Of note, alterations in ceramides have been previously associated with lymph node-positive PDAC (25). To ensure that our observed alterations in sphingolipids in cachectic patients were not simply a reflection of gross metastasis, we repeated our analysis excluding the six patients in the cachectic group with metastatic (M1) disease.

All three previously identified differences in plasma ceramide content remained (Supplemental Figures 3A and 3B), suggesting that the decreases in blood sphingolipid content of cachectic PDAC patients are not simply a consequence of metastatic disease.

**Table 1.**
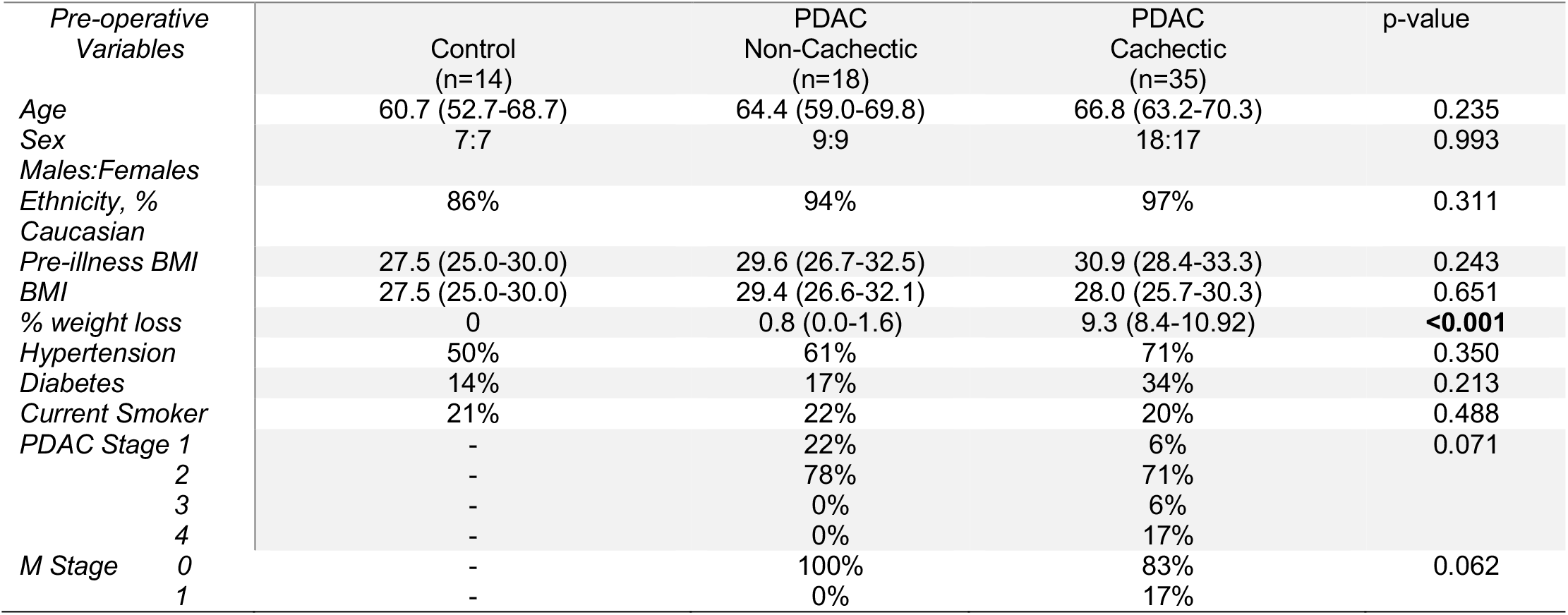
Clinical Characteristics of the combined cohort.

### Plasma ceramides as biomarkers for cancer cachexia

To determine whether alterations in circulating sphingolipid content could serve as a biomarker of PDAC-induced cachexia, the Biomarker Analysis feature of MetaboAnalyst was used to perform Receiver Operating Characteristic (ROC) analysis. This analysis tested the ability of a given sphingolipid to distinguish cachectic from non-cachectic cancer patients, thus excluding non-cancer controls. Of our targeted metabolites, only C24 ceramide was able to significantly separate cachectic patients, with an area under the ROC curve (AUROC) of 0.730 (95% CI 0.598-0.858, p= 0.0074, Figure 3A).

**Figure 3.**
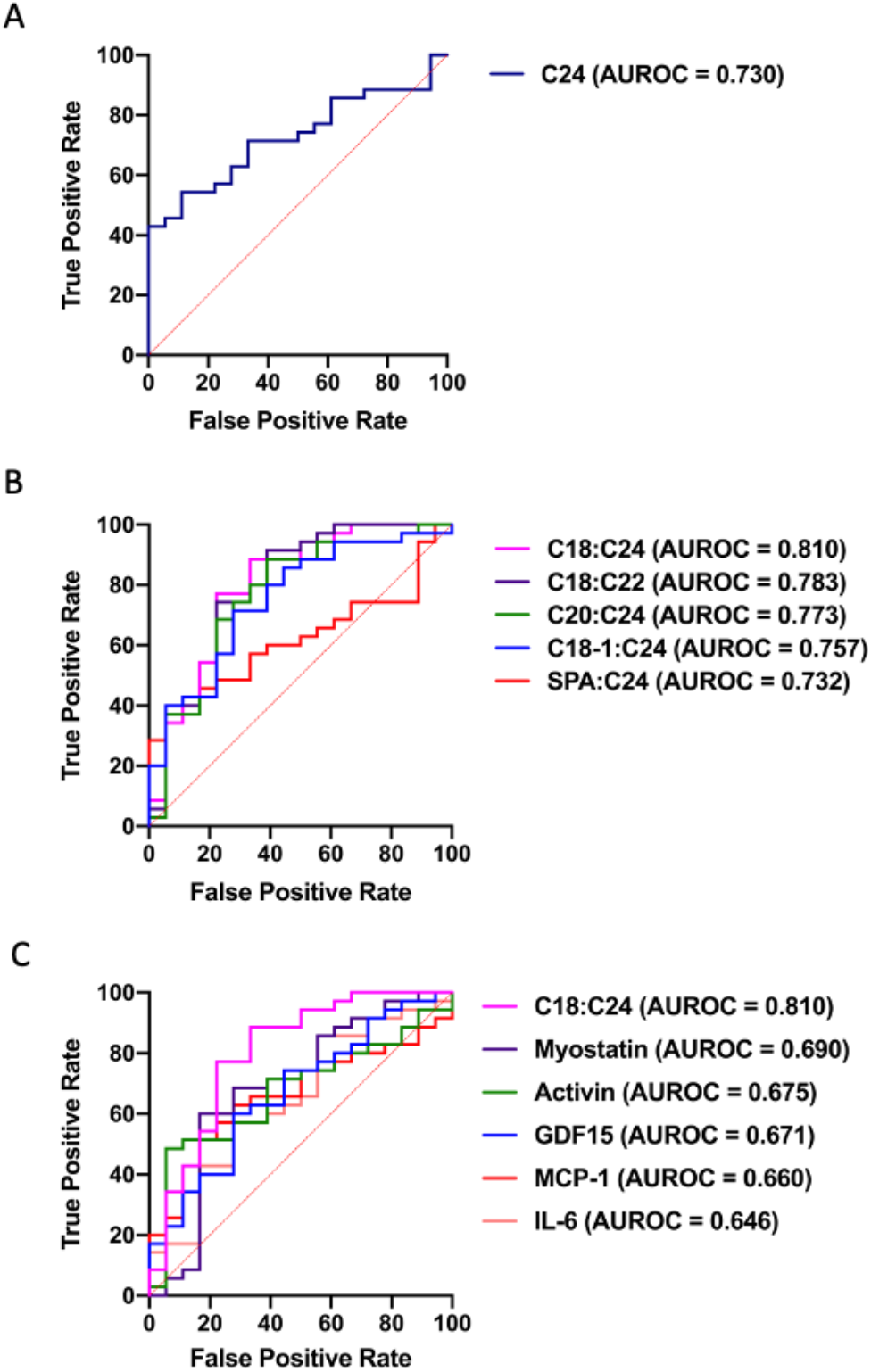
C24, ceramide ratios, and classical circulating factors as biomarkers of cancer cachexia. (A) ROC curve for distinguishing non-cachectic from cachectic PDAC patients by C24. (B) ROC curves for distinguishing non-cachectic from cachectic PDAC patients by ratios of sphingolipids (C) ROC curve for C18:C24 distinguishing non-cachectic from cachectic PDAC patients compared to ROC curves for other previously proposed biomarkers of cancer cachexia. ROC curves are displayed as sensitivity vs. (1-specificity) value. n=18 noncachectic PDAC patients, n=35 cachectic PDAC patients.

Because a ratio of plasma ceramides has been previously established as an indicator of mortality risk marker in individuals with cardiovascular disease (26), we posited that a ratio of plasma ceramides might serve as a more effective marker of cancer cachexia than a single ceramide. Indeed, 18 ratios of plasma sphingolipids were able to identify cachectic PDAC patients. Five of these ratios had AUROCs greater than that of C24 alone, with C18:C24 exhibiting the highest AUROC (Table 2, Figure 3B).

**Table 2.** Plasma ceramide ratios are able to distinguish cachectic from non-cachectic PDAC patients.

|  | Non-Cachectic (n=18) | Cachectic (n=35) | AUROC (95% CI) | p-value |
| --- | --- | --- | --- | --- |
| C18:C24 | 0.0933 ± 0.0387 | 0.2009 ± 0.0467 | 0.810 (0.679-0.921) | 0.0087 |
| C18:C22 | 0.2236 ± 0.0495 | 0.3757 ± 0.0546 | 0.783 (0.633-0.910) | 0.0075 |
| C20:C24 | 0.1042 ± 0.0404 | 0.1923 ± 0.0478 | 0.773 (0.630-0.903) | 0.0196 |
| C18-1:C24 | 0.0021 ± 0.0005 | 0.0064 ± 0.0015 | 0.757 (0.615-0.8) | 0.0048 |
| SPA:C24 | 0.0028 ± 0.0003 | 0.0042 ± 0.0005 | 0.732 (0.593-0.863) | 0.0098 |
| C24 alone | 3.097 ± 0.1577 | 2.474 ± 0.1627 | 0.730 (0.598-0.858) | 0.0074 |

Mean ± SEM, AUROC (95% CI of the mean), p values are from ROC analysis.

### C18:C24 values are affected by neo-adjuvant treatment

In our previous biomarker study, we showed that the association between MCP-1 and cancer cachexia was lost in patients who had received neoadjuvant treatment. To determine if C18:C24 ratio was similarly affected, we selected a cohort of cachectic and non-cachectic PDAC patients who had received chemotherapy with or without radiotherapy prior to their attempted tumor resection and plasma collection (clinical data appear in Supplemental Table 3). Similar to our findings with MCP-1, C18:C24 was unable to distinguish treated cachectic patients from non-cachectic patients (AUROC = 0.579, p = 0.949, Supplemental Figure 4). Such findings highlight the potential limitations of chemotherapy and/or radiotherapy on cachexia biomarker studies.

**Figure 4.**
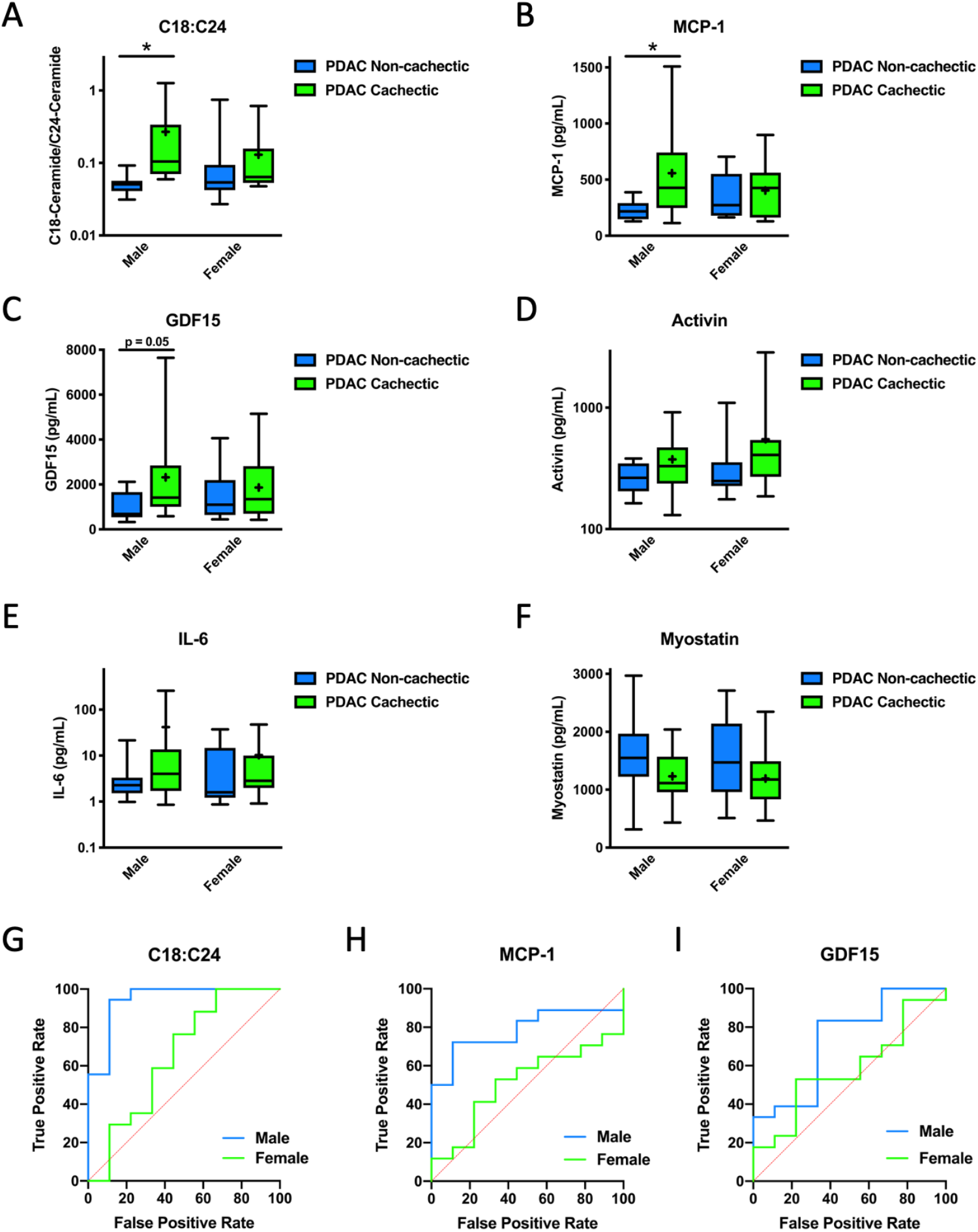
C18:C24 and other biomarkers of cancer cachexia are sexually dimorphic. (A) C18:C24 and (B) MCP-1 are increased in cachectic male PDAC patients, but not female patients, and (C) GDF15 tends to be increased in cachectic males but not females. Circulating levels of (D) activin, (E) IL-6, or (F) myostatin were not increased in cachectic male or female patients. (G-I) Clear differences exist between ROC curves for males and females. In box-and-whiskers plots, the box is the 25^th^/75^th^ percentile with minimum-to-maximum whiskers. The line represents the median with the + representing the mean. ROC curves are displayed as sensitivity vs. (1-specificity) value. n=18 non-cachectic PDAC patients, n=35 cachectic PDAC patients. * represents significantly different from control of the same sex, p < 0.05 by two-way ANOVA with sex and cachexia as variables.

### C18:C24 outperforms previously proposed biomarkers of cancer cachexia

To test the robustness of C18:C24 as a cachexia biomarker, we performed a head-to-head comparison with a number of other circulating factors that have been proposed as biomarkers of cancer cachexia (16, 27-29). High-sensitivity ELISAs for IL-6, activin, Growth Differentiation Factor 15 (GDF15), myostatin, and MCP-1 were performed from the same plasma samples used in our metabolomic analysis. Similar to previous findings (15, 16), IL-6 was not able to distinguish non-cachectic and cachectic PDAC patients. Likewise, myostatin or activin levels were not different between groups. Significantly, C18:C24 exhibited a greater AUROC compared to each of the other proposed biomarkers that we tested (Table 3 and Figure 3C).

**Table 3.** ROC Curve Analysis of Proposed Cachexia Biomarkers.

|  | Non-Cachectic (n=18) | Cachectic (n=35) | AUROC (95% CI) | p-value |
| --- | --- | --- | --- | --- |
| C18:C24 | 0.0933 ± 0.0387 | 0.201 ± 0.0467 | 0.810 (0.679-0.921) | **0.0087** |
| MCP-1 (pg/mL) | 290.4 ± 38.9 | 482.5 ± 57.9 | 0.660 (0.513-0.797) | **0.0410** |
| GDF15 (pg/mL) | 1233 ± 235 | 2097 ± 309 | 0.671 (0.489-0.800) | **0.0431** |
| Activin (pg/mL) | 314.7 ± 48.5 | 456.7 ± 77.2 | 0.675 (0.521-0.832) | 0.0868 |
| IL-6 (pg/mL) | 6.36 ± 2.44 | 26.47 ± 10.76 | 0.646 (0.470-0.818) | 0.1259 |
| Myostatin (pg/mL) | 1647 ± 162 | 1213 ± 77 | 0.690 (0.505-0.856) | 0.1293 |

Mean ± SEM, AUROC (95% CI of the mean), p values are from ROC analysis.

### C18:C24 and other proposed biomarkers are sexually dimorphic

Since recent studies have pointed to potential sex differences in cancer cachexia, we sought to determine if a sexual dimorphism existed with our newly identified C18:C24 biomarker. Results showed that plasma levels of C18:C24 associated with PDAC-induced cachexia, but notably only in male patients (Figure 4A).

We observed that a similar dimorphism occurred with MCP-1 (Figure 4B) and GDF15 (Figure 4C). In contrast, plasma levels of activin (Figure 4D), IL-6 (Figure 4E), and myostatin (Figure 4F) were not different between male and female cachectic and non-cachectic patients. By ROC analysis, C18:C24, MCP-1, and GDF15 were able to separate male cachectic PDAC patients from non-cachectic patients, but not female patients. C18:C24 again exhibited the highest AUROC (Table 4). For comparison purposes, ROC curves for both males and females are overlaid for C18:C24 (Figure 4G), and MCP-1 (Figure 4H), and GDF15 (Figure 4I).

## Discussion

Our results reported here identify plasma ceramide ratios, and particularly C18:C24, as a potential biomarker for PDAC-induced cachexia. Further, our results indicate that patient characteristics such a sex should be considered when new biomarkers are assessed.

Our data are consistent with previous work in which decreases in C24-ceramide and S1P were found to be part of a metabolic signature capable of distinguishing patients with PDAC from patients with other GI disorders (30). While this study did not consider weight loss, it is tempting to speculate that the high incidence of weight loss in PDAC patients may have contributed to this finding. Alterations in plasma ceramide levels have also been reported with obesity and type 2 diabetes (31). Although we did not find a linear relationship between a patient’s body mass index and their C18:C24 ratio (data not shown), future biomarker studies should consider adiposity as factor for such an analysis. Similar consideration should be given to diabetes, which is also common in PDAC patients.

Several caveats should be noted in our study. First, while we used the international consensus definition for cancer cachexia to stratify our patients (1), recent work suggests that weight loss of 5% may not be associated with decreased survival in PDAC patients (8, 15). Furthermore, because our study was performed at a single institution, our results reflects the patient demographics of that site, which were disproportionally non-Hispanic Caucasians. Thus, our data are not reflective of the more diverse population that develops PDAC-induced cachexia. Moreover, our dataset is limited to PDAC patients undergoing a surgical procedure and thus not representative of the majority of patients diagnosed with advanced PDAC. Thus, we do not know if the C18:C24 ratio identified in cachectic patients with early stage PDAC would be generalizable to patients with more advance disease or cachexia induced by other cancers. Such factors need to be addressed in subsequent studies.

Our data also uncovered an interesting finding related to differences in circulating C18:C24 between cachectic and non-cachectic PDAC patients that appear to be primarily driven by differences in male patients. It is increasingly recognized that in addition to varying by age, metabolite profiles also differ between sexes (32). However, we note that this sexual dimorphism is not specific to ceramides, as similar differences occurred in MCP-1 and GDF15. While it is certainly possible that this apparent sex difference in cachexia-associated biomarkers is tied to basal differences in sphingolipid levels or the accuracy of self-reported weight loss between males and females (33-35), our data make a compelling argument for the necessity of considering sex as a biological variable in future efforts identify a biomarker of cancer cachexia.

## Methods

### Biobank

Patients 18 years of age and older undergoing an abdominal operation for pancreatic cancer or other benign conditions were eligible for enrollment into the Ohio State Pancreatic Cancer Cachexia tissue registry. A detailed patient history as well as height and weight measurements were taken at the preoperative surgical clinic visit to determine the history of weight loss. All other variables were abstracted from the pre-operative history and physical, as well as the electronic medical record. Consistent with the international definition, cachexia was defined as more than 5% loss of body weight over the previous 6 months (1). Patients with >2% but <5% loss of pre-illness body weight and were confirmed to have a body mass index (BMI) of less than 20 kg/m^2^ were considered cachectic. Because not every enrolled patient had a CT scan available for analysis, patients were not assessed for cachexia based upon muscle volume.

### Preparation of plasma

For patients who elected to contribute to the biobank, approximately 30 cc peripheral blood was collected intraoperatively following induction of anesthesia in heparinized tubes. Following centrifugation at 500 g for 10 minutes, plasma was aliquoted and stored at −80 °C until shipping.

### Study design

Our initial cross-sectional study design involved choosing three patient cohorts for analysis from our institutional tissue bank: 1) control patients without active cancer or inflammatory conditions undergoing elective abdominal operations for a variety of diagnoses and with no recent history of weight loss; 2) weight-stable (non-cachectic) patients with pathology-confirmed pancreatic adenocarcinoma; and 3) cachectic patients with pathology-confirmed pancreatic adenocarcinoma. Groups consisted of five males and five females. To the best of our ability, patients were matched based on age and body mass index. We elected to limit our cachectic patient cohort to patients with weight loss between 5 and 15% weight loss to avoid including patients with refractory cachexia (1). Following our initial results, additional samples were analyzed to confirm our initial findings. In our second cohort, we allowed patients with weight loss of less than 5% to be included in our study. We did not pre-register our experimental plan.

### Metabolomic sample processing and analysis

Frozen plasma samples were shipped to the Mayo Clinic for analysis by liquid chromatography followed by tandem mass spectrometry to determine absolute metabolite concentrations were based on reference standards, with the exception of tricarboxylic acid cycle intermediates, which were analyzed by gas chromatography. Samples were run in random order to minimize drift, and technical staff were blinded to the sample groups.

### Enzyme-linked immunosorbent assay (ELISA)

Plasma levels of IL-6, activin, GDF15, myostatin, and MCP-1 were determined by ELISA. Kits for myostatin, GDF15, and activin were purchased from R&D Systems, while high sensitivity IL-6 and MCP-1 kits were purchased from eBioscience. Samples were analyzed in duplicate and results averaged.

### Statistics

For comparisons of clinical data with three groups, One-way Analysis of Variance (ANOVA) was used for continuous variables, and Chi Square was used for binary or nominal variables. Comparisons of clinical data with two groups were performed using Student’s t-tests for continuous variables and Chi Square for binary or nominal variables.

Metabolite analysis was performed using MetaboAnalyst (36). Samples were normalized to the median of each metabolite analyzed, and then data were transformed by cube-root transformation due to the substantial number of near-zero concentrations of ceramides (37). Pareto scaling was then performed. Differences between groups were then assessed using one-way ANOVA with Fisher’s LSD post-hoc using a false discovery rate of 0.05. Heatmaps were generated in MetaboAnalyst using Euclidean distant measurements, auto-scaled by features, and colored according to the normalized values. Graphs were created in GraphPad Prism 8.3.

Ability to discriminate between pancreatic cancer patients with and without cachexia was determined by Receiver Operating Characteristic (ROC) analysis using the Biomarker Analysis feature of MetaboAnalyst 4.0 (36). Area Under the Receiver Operating Characteristic Curve (AUROC) is reported as a measure of a biomarker’s potential as a circulating marker of pancreatic cancer-induced cachexia. MetaboAnalyst 4.0 was utilized for this analysis, with data log transformed and auto-scaled as suggested by Chong et al. (36). However, similar results were obtained without transformation and scaling. Graphical depictions of ROC curves were generated using GraphPad Prism and may vary slightly from those generated by MetaboAnalyst 4.0, as they lack normalization. However, AUROC and p values were similar by both methods. The ROC curves are displayed as sensitivity vs. (1-specificity) value.

Differences in levels of circulating factors between sexes were assessed by two-way ANOVAs with sex and cachexia as factors on log-transformed data, followed by Sidak’s multiple comparisons test.

### Study approval

All aspects of the study were approved by The Ohio State University Institutional Review Board. Written informed consent was obtained from each patient prior to enrollment in the tissue bank.

## Data Availability

Requests for primary data will be considered after consultation with the involved IRBs.

## Author Contributions

DCG and EET designed the study; PVR and EET conducted experiments; JMC and EET analyzed data; MED, CRS, DCE, and TMW provided and organized patient samples; JMC, DCG, EET wrote the manuscript; all authors edited and approved a final version of the manuscript.

## Acknowledgements

This publication was made possible by a pilot grant from the Mayo Clinic Metabolomics Resource Core through grant number U24DK100469 from the National Institute of Diabetes and Digestive and Kidney Diseases, which originates from the National Institutes of Health Director’s Common Fund. The authors would like to thank the Mayo Clinic Metabolomics Core, and particularly Xuan-Mai T. Petterson, for sample processing and coordination. Additional support was provided by the National Cancer Institute P30 CA016058 (The Ohio State University) a National Institute of Cancer R01 CA180057 (DCG), and a Weiss Postdoctoral Fellowship (EET).

